# Discerning Social Determinants of Health Associated with Blood Pressure Control of Hypertension for Patients at Murang’a County Referral Hospital, Kenya Synthesizing a Hypertensive Patient Profile for SDOH at Risk

**DOI:** 10.1101/2025.09.04.25335098

**Authors:** Okubastion Tekeste Okube, Vincencia Aketch, Nerry Kitur, Josphat Njoroge, Norah Anne Mogute Oyagi, Lee Presley Gary

## Abstract

Our study investigated the link between blood pressure control and the Social Determinants of Health of 142 adult hypertensive patients, treated at Murang’a County Referral Hospital, Kenya, during November 2023 while receiving blood pressure interventions. Blood pressure readings were obtained from hospital records. A systematic random sampling technique was used for recruiting participants. Quantitative data was collected through a semi structured questionnaire to assess categorical variables: demographics, income, education, employment, health literacy, and access to healthcare. Data was analyzed using Statistical Package for Social Sciences software version 22.

Our research revealed that, with the availability of hypertensive therapies, individual blood pressure rates remain suboptimal for the participant population. We found that only 43% (61) achieved targeted blood pressure levels at less than 140/90 mm Hg. Concurrently, we monitored significant determinants of blood pressure control, including advanced age, illiteracy, less frequency of hospital visits, lack of access and affordability of healthcare services, and lack of psychosocial support system.

Our research confirmed that adult hypertension and uncontrolled blood pressure together remain leading contributors to cardiovascular issues in Murang’a community, and Social Determinants of Health were at serious risk for unsuspecting individuals, each playing a critical role with influencing individual health outcomes.

## 1. INTRODUCTION

The World Health Organization (WHO, 2013), defines hypertension (HTN) or uncontrolled blood pressure (BP), as a systolic and/or diastolic blood pressure equal to or above 140/90 mmHg respectively. Uncontrolled blood pressure stands as a major risk factor for several cardiovascular events, including ischemic heart disease, myocardial infarction, hemorrhagic stroke, and heart failure (Lippi et al., 2020). Despite remarkable progress in the field of healthcare, plus measures put in place to prevent and control cardiovascular disease (CVD), hypertension remains a significant public health problem worldwide. In 2019, the global prevalence of hypertension in adults aged 30–79 years was 32% in women and 34% in men(Nguyen & Chow, 2021). However, only 54% of adults with the condition are diagnosed, 42% receive treatment, and only 21% have their blood pressure controlled (WHO, 2023a).

In Sub-Saharan Africa (SSA), WHO estimates that 46% of adults, aged above 25 years, have some degree of hypertension (Yoruk et al., 2018). According to Zhou et al. (2021a), the prevalence of adult hypertension in Sub-Saharan African countries was 48% in women and 34% in men in 2019. However, in the less developed countries, including SSA, it is estimated that 46% of the people with hypertension are not even aware that they have this condition (WHO, 2023-a).

In the less developed nations, diagnosis and treatment of hypertension are often delayed due to a a lack of awareness and screening, leading to increased risk of complications and mortality (Dhungana et al., 2016). In SSA, screening, diagnosis, and treatment are inadequate and a recent study found that 40% of individuals with the condition in East and West Africa were unaware of their status (Ibekwe, 2015). In Kenya, reports have given variable estimates for the prevalence of hypertension: including 44.7% (Ondieki et *al.,* 2021) and 20.8% (Mecha et al., 2020). Correspondingly, in Kenya, only 49% of adults with the condition are diagnosed, with 21% treated, and only 8% have their blood pressure controlled (WHO, 2023b). These stats help to underscore the necessity for improved awareness, detection and management of adult hypertension.

Uncontrolled blood pressure is the leading cause of death globally, attributed to 10.4 million deaths in 2017 (Stanaway et al., 2018). Low- and middle-income countries (LMICs) suffer the most, with less than 20% of individuals achieving adequate control, while hypertensive-related cardiovascular deaths have been rising over the past 3 decades (Ferdinand and Brown, 2021; Ogunniyi et al., 2021). More than 75% of deaths caused by cardiovascular diseases occur in the LMICs, attributed to undiagnosed, or to untreated, or inadequately treated hypertension sources (WHO, 2019). For many people in such countries, detection is often late in the course of the disease, and people die at a younger age from CVDs often in their most productive years (WHO, 2019).

Likewise, in Kenya, cardiovascular diseases are a significant public health burden, accounting for approximately 26% of all Non-Communicable Diseases (NCD)-related deaths (Wekesah et al., 2020). Moreover, cardiovascular-related complications required long-term hospitalization, putting families and individuals into financial crisis due to out-of-pocket payment. At worst catastrophic health expenses from cardiovascular issues disproportionately affect low-income communities, perpetuating suffering, and deepening and expanding socioeconomic disparities (Gheorghe et al., 2018).

In addition to lifestyle risk factors, like a poor diet, physical inactivity, alcohol and tobacco use, the development and management of hypertension are significantly influenced by an individual’s Social Determinants of Health (SDOH). Overall, hypertension is significantly impacted by the SDOH characteristics, such as age, gender, education, income, employment and job stability, access to health care and social support networks (Virani et al., 2021; Wang, Kho, & French, 2020). The burden of hypertension and related cardiovascular issues fall disproportionately on those from socially disadvantaged backgrounds (Benjamin et al., 2019; Minhas et al., 2023).

Because they are more likely to be exposed to suspect conditions and have less access to good screening and initial treatment than people with favorable socioeconomic standing, vulnerable and socially disadvantaged individuals suffer from hypertension more often and die sooner from cardiovascular diseases (Havranek et al., 2015). Poor SDOH can shape an individual’s unhealthy behaviors, fostering unhealthy practices that directly impact stress levels, leading to a heightened sympathetic activity, key markers of inflammation, and enhanced susceptibility to uncontrolled hypertension and then cardiovascular issues (Lurbe and Ingelfinger, 2021). Limited access to affordable, high-quality health care compounds these challenges, hindering timely diagnosis and treatment, and leading to an accelerated progression of problems with high morbidity and mortality (Gheorghe et al., 2018). Indeed, cardiovascular events, including heart attacks, strokes, and mental fitness, are significantly influenced by SDOH (Jilani et al., 2021).

Reducing and controlling hypertension is one of WHO’s worldwide goals for the prevention of non-communicable diseases (NCD Risk Factor Collaboration, 2021). Despite significant efforts to address the challenges associated with blood pressure control, social determinants remain an issue of concern with disparities in accessing the quality of healthcare services (Zhou B et al., 2021a; Zhou B et al., 2021b). Understanding the relationship between SDOH and adequate blood pressure control is necessary to design appropriate interventions and key implementation strategies to improve access to quality healthcare and reduce the burden of any cardiovascular diseases in the lesser developed nations. As such, this research study aimed at discerning the latent association between SDOH and blood pressure intervention among adult patients with hypertension receiving treatment at the Murang’a County Referral Hospital in Kenya.

## 2. LITERATURE SUMMARY

Globally, hypertension is a primary cause of morbidity and mortality. The global prevalence of this condition in adults aged 18 years and above was 24% for men and 20% in women in 2015 (Zhou et al., 2017). Sadly, hypertension is even more common in Sub-Saharan African countries as high as 46% of adults aged above 25 years (Yoruk et al., 2018) are affected. Moreover, in the SSA, detection rates are exceptionally low due to inadequate screening and to uptakes for blood pressure checks, mixed with low medical adherence to treatment (Sorato, 2021). For Kenya, the prevalence of adult hypertension ranges from 20.8% (Mecha et al., 2020) to 44.7% (Ondieki et *al.,* 2021), while blood pressure care remains low at 33.4% (Mbau et al., 2022).

## 3. STUDY METHODS and MATERIALS

### 3.1 Study Area

Our research study was carried out at Murang’a County Referral Hospital, located in Murang’a town, Kenya during November 2023. The hospital was founded in the early 1950s, and currently, it is a Level - 5 hospital and is the largest public hospital in Murang’a County with bed capacity of 317. The hospital offers inpatient and out-patient services including intensive care, maternity, renal, psychiatry, pediatrics, hypertension, plus a diabetic clinic and other specialized clinics.

A study of patients at Murang’a County Referral Hospital showed that uncontrolled levels of their blood pressure were as high as 59.3% (Eunice et al., 2018). Murang’a County is one of areas where the number of hypertensive patients has been rising steadily but management of care remains inadequate. Many people living with hypertension in Murang’a County remain unaware of their condition, thus putting themselves at the risk of developing further medical complications, ultimately death.

### 3.2 Study Design, Participants and Sampling Techniques

A health facility-based cross-sectional study design was carried out among adult patients with known hypertension attending the Referral Hospital. A systematic random sampling technique was used in recruiting the participants. As an inclusion criterion, all hypertensive patients who attended Murang’a County Referral Hospital during the time of the study were included, while the exclusion criteria covered hypertensive patients who declined to give consent for the study and those who were critically ill and could not communicate on their own. Patients with known hypertension who met the inclusion criteria were considered eligible for the study. According to the hospital’s records, approximately 324 patients with hypertension attended the MCRH every month. A sampling interval of two was determined by dividing the target population (n=324) by the calculated sample size (142). Thus every 2nd patient who attended the hospital during the study period was selected to participate in the study until the desired sample size was achieved.

### 3.3 Sample Size Determination

The Fisher’s formula was used to determine sample size

n=[Z^2^ (p)(q)]\d^2^
n= sample size
Z: statistic for a level of confidence and for a level of confidence of 96% Z value is 1.96.
P: the proportion of population with the desired characteristics was taken at 20.8% from a study done in Kenya (Mecha et al., 2020).
q: (1-p)-the proportion of population without the desired characteristics.
d: margin of error taken as 5% (0.05).
n= {1.96^2^ ×0.208×0.792}/0.05^2^, n= 253.

Since the target population was less than 10,000 in one month period, the sample size was further adjusted using: nf= n/1+n/N. where, nf= population less than 10,000.

N= total population during the time of study in MCRH, patients’ population of 324.
n= calculated sample size =253.
Calculating, nf = 253/1+ 253/ 324 = 142.
Therefore, the sample size for this study was 142.

### 3.4 Data Collection Tools

Semi-structured, printed questionnaire was used to collect quantitative data that included socio-demographic characteristics, social support networks, and access to healthcare. The BP value was extracted from the patients’ clinical records. The level of BP values was categorized as controlled less than 140/90 mmHg, and uncontrolled as greater than or equal to 140/90 mmHg (WHO, 2020). Pre-testing of the data collection tools was done on 15 randomly selected patients.

### 3.5 Ethics Approval

Initial approval to carry out our study in Murang’a County Referral Hospital was obtained from The Catholic University of Eastern Africa’s School of Nursing. Our research study protocol was approved by the Kenyatta National Hospital and the University of Nairobi Research and Ethics Committee with assigned approval No: UP477/07/2023. Our Research Permit was granted by the National Commission for Science, Technology, and Innovation and was assigned License No: NACOSTI /P/23/30842 with endorsement from the hospital administration.

The research program and procedures were explained verbally to the individuals who were willing and able to voluntarily participate in the study. Willing participants signed consent papers before inclusion into the study. Strict privacy was maintained throughout the study, and to maintain confidentiality, personal identifications of the participants were not included in the questionnaire. All filled questionnaires were kept confidential and protected, and they were only accessed by the principal researcher and authorized research personnel.

### 3.6 Data Analysis

Daily verification of all collected data was routinely conducted by the principal researcher with the help of the research assistants to ensure completeness of the collected data. Data analysis was done using the Statistical Package for Social Science (SPSS) with software (version 22.0). Descriptive statistics were expressed as frequencies and percentages. The relationship between independent and dependent variables for categorical variables was determined using *chi-square* test. A *p-value* less than 0.05 was considered as statistically significant.

## 4. RESULTS

### 4.1 Socio-Demographic Characteristics

Table 1 displays the socio demographic characteristics of the participants. Approximately one third (34.5%) of the respondents had age range between 46-55 years and 56-65 years. Majority were male (58.5%), married (51.4%), Protestants (52.8%), residents in the rural areas (56.3%) and reported having monthly income of less or equal to Ksh.10, 000 (62.7%). Nearly one-third of the respondents attained primary level of education (39.4%) and were self-employed (35.2%).

**Table 1:**
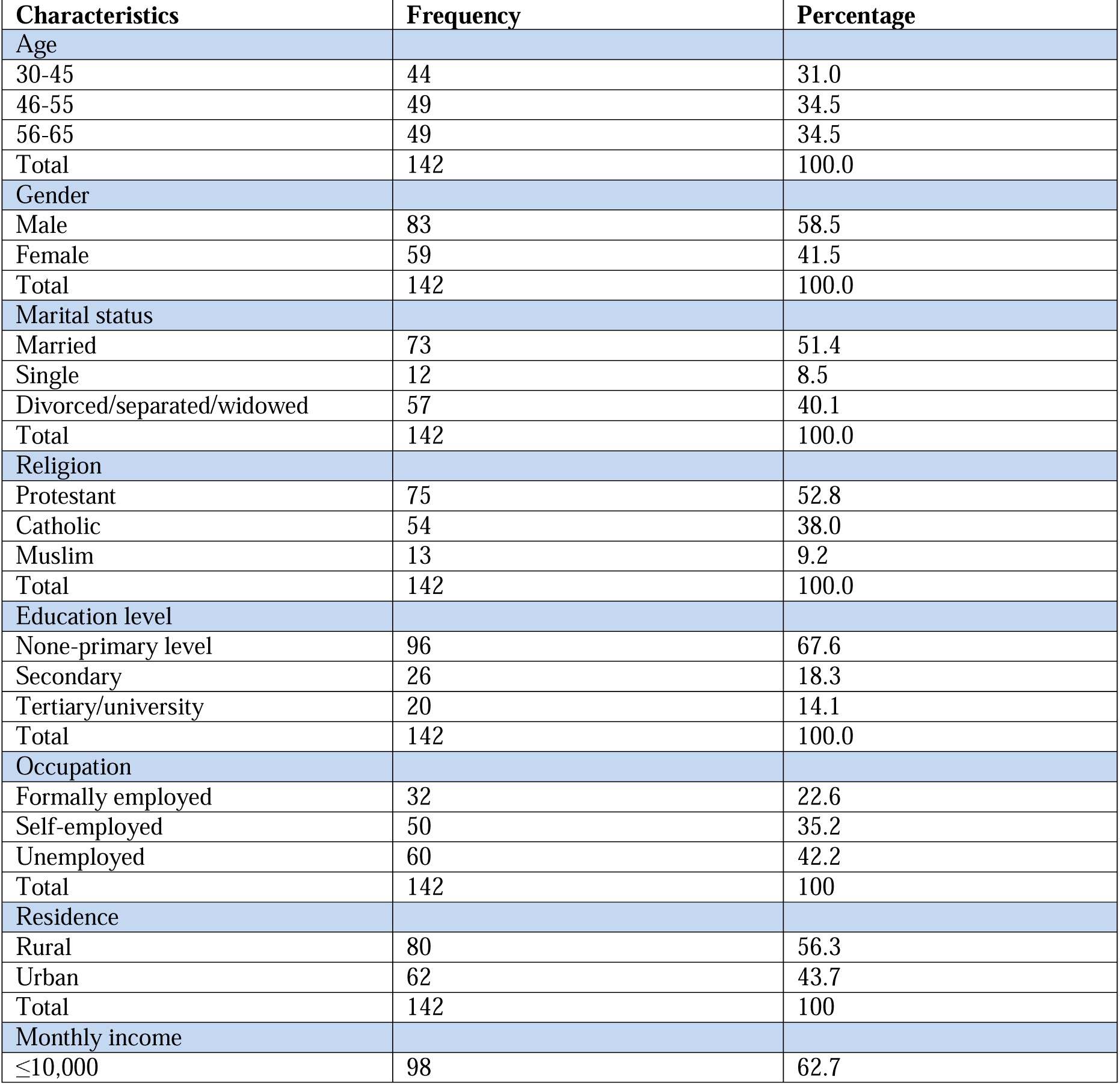
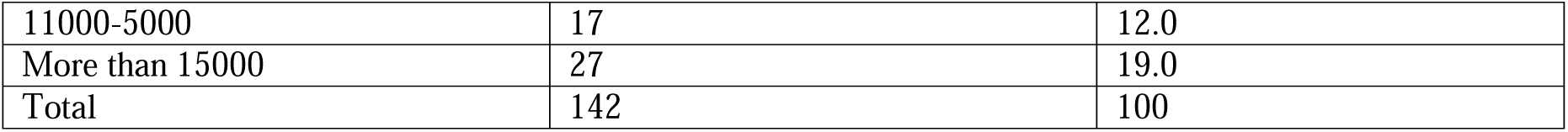
Socio-Demographic Characteristics of the Respondents.

| Characteristics | Frequency | Percentage |
| --- | --- | --- |
| **Age** |  |  |
| 30-45 | 44 | 31.0 |
| 46-55 | 49 | 34.5 |
| 56-65 | 49 | 34.5 |
| Total | 142 | 100.0 |
| **Gender** |  |  |
| Male | 83 | 58.5 |
| Female | 59 | 41.5 |
| Total | 142 | 100.0 |
| **Marital status** |  |  |
| Married | 73 | 51.4 |
| Single | 12 | 8.5 |
| Divorced/separated/widowed | 57 | 40.1 |
| Total | 142 | 100.0 |
| **Religion** |  |  |
| Protestant | 75 | 52.8 |
| Catholic | 54 | 38.0 |
| Muslim | 13 | 9.2 |
| Total | 142 | 100.0 |
| **Education level** |  |  |
| None-primary level | 96 | 67.6 |
| Secondary | 26 | 18.3 |
| Tertiary/university | 20 | 14.1 |
| Total | 142 | 100.0 |
| **Occupation** |  |  |
| Formally employed | 32 | 22.6 |
| Self-employed | 50 | 35.2 |
| Unemployed | 60 | 42.2 |
| Total | 142 | 100 |
| **Residence** |  |  |
| Rural | 80 | 56.3 |
| Urban | 62 | 43.7 |
| Total | 142 | 100 |
| **Monthly income** |  |  |
| ≤10,000 | 98 | 62.7 |
| 11000-5000 | 17 | 12.0 |
| More than 15000 | 27 | 19.0 |
| Total | 142 | 100 |

Of the 142 respondents, 35.9% covered less than 1 km and 37.3% covered 1-3 km to visit the Referral Hospital, while approximately 26.8% covered more than 3 km to the nearest hospital. Majority (62.7%) of the respondents used tarmac road to visit the nearest hospital. A higher proportion (45.8%) of the respondents travelled to the nearest hospital by foot, and 37.3% used public transport.

Most, (76.1%) of the respondents reported being able to afford a means of transport to visit the hospital (76.1%). Majority of the respondents (52.8%) visited the hospital more than once a month while 47.2% visited only once in a month. Of the respondents 57.7% did not access the medical professionals in time. On affordability of services provided, 68.3% responded that the services were not easily affordable while 31.7% responded services were affordable. Of all the respondents, 51.4% paid their medical bills by cash while 48.6% paid using insurance.

**Table 2:** Accessibility to Healthcare.

| Characteristics | Frequency(n) | Percentage (%) |
| --- | --- | --- |
| **Distance** |  |  |
| Less than 1km | 51 | 35.9 |
| 1-3 km | 53 | 37.3 |
| More than 3km | 38 | 26.8 |
| Total | 142 | 100 |
| **Type of road** |  |  |
| Murram | 53 | 37.3 |
| Tarmac | 89 | 62.7 |
| Total | 142 | 100 |
| **Transport means** |  |  |
| Foot | 65 | 45.8 |
| Private | 24 | 16.9 |
| Public | 53 | 37.3 |
| Total | 142 | 100 |
| **Is the means of transport affordable** |  |  |
| No | 34 | 23.9 |
| Yes | 108 | 76.1 |
| Total | 142 | 100 |
| **Visits to hospital** |  |  |
| Once | 67 | 47.2 |
| More than once | 75 | 52.8 |
| Total | 142 | 100 |
| **Access** |  |  |
| No | 82 | 57.7 |
| Yes | 60 | 42.3 |
| Total | 142 | 100 |
| **Services** |  |  |
| No | 97 | 68.3 |
| Yes | 45 | 31.7 |
| Total | 142 | 100 |
| **Medical bills** |  |  |
| Cash | 73 | 51.4 |
| Insurance | 69 | 48.6 |
| Total | 142 | 100 |

### 4.3 Availability of Social Support Network

On availability of social support networks, a majority (76.1%) responded that they had friends and family visit with them when sick while others (23.9% responded that they did not have both contacts while sick. Majority of the respondents (75.4%) had emotional support while others (24.6%) did not have emotional support. On financial support, 64.8% had good financial support and 35.2% did not have the financial support. With problem solving and serious decision-making challenges, 63.4% had people to help them solve problems, and 36.6% did not receive or found available support.

**Table 3:**
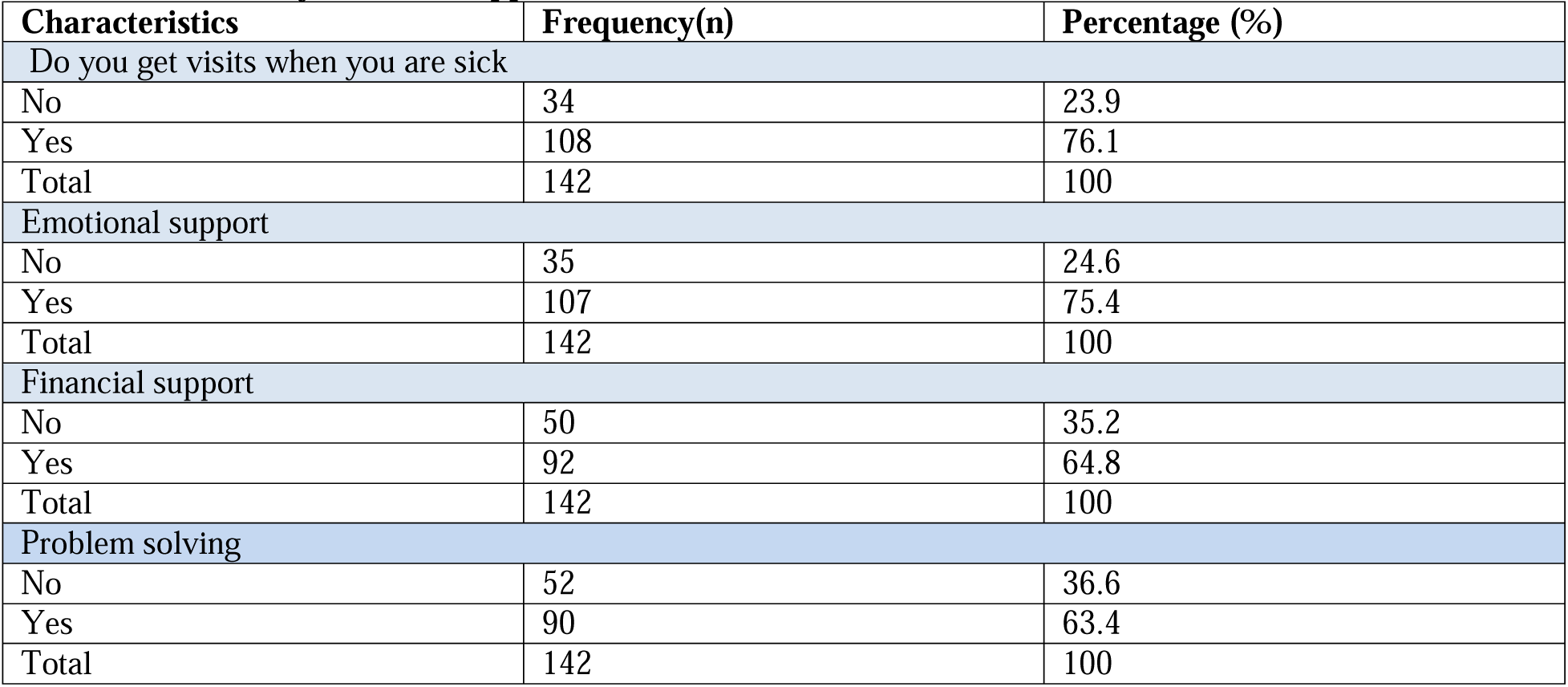
Availability of Social Support Networks.

### 4.4 Level of Blood Pressure Among the Respondents

Figure 1 presents the proportion of respondents with controlled and uncontrolled blood pressure. Most (57%) of the respondents had uncontrolled levels.

**Figure 1:**
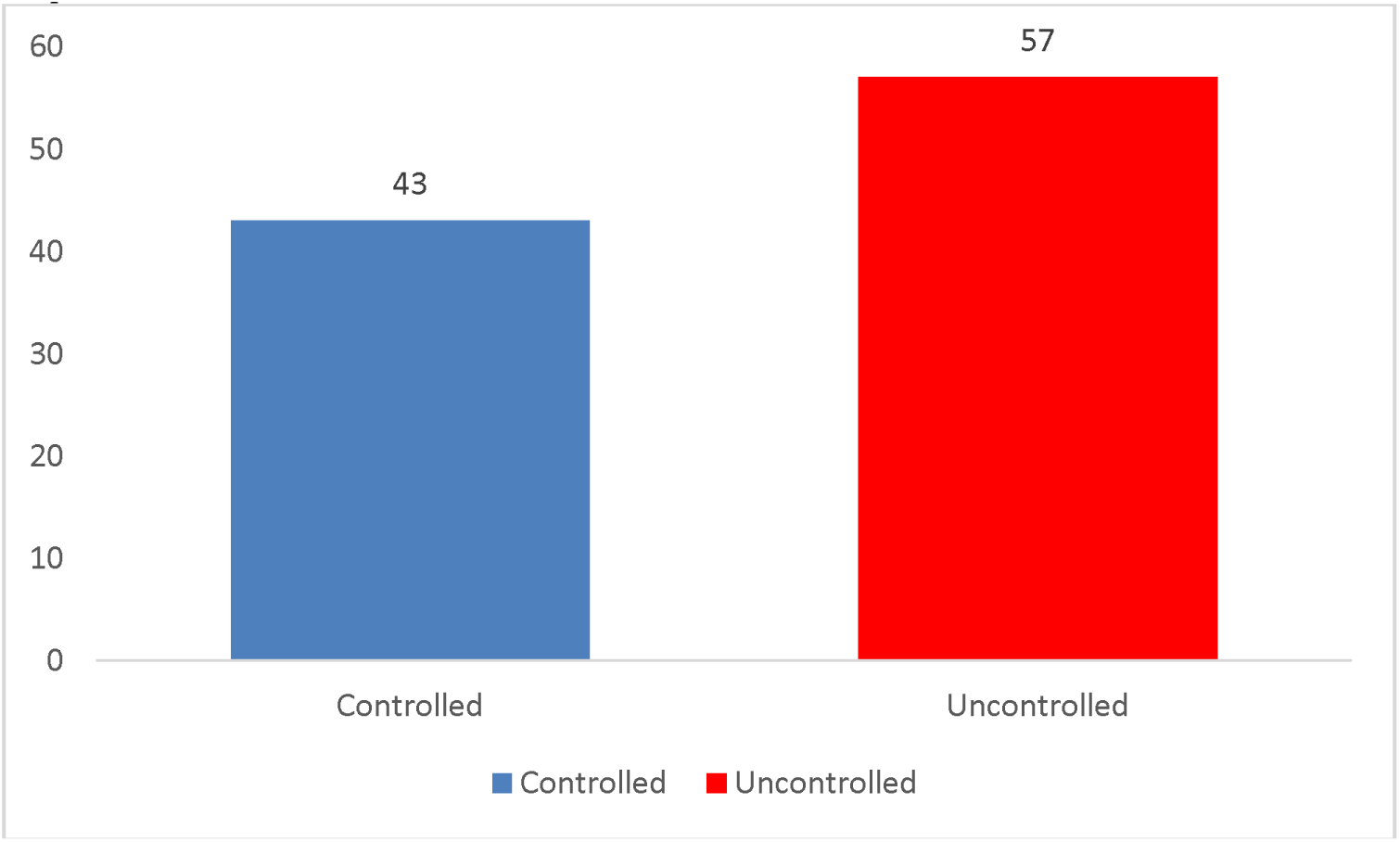
**Level of Blood Pressure Among the Respondents.**

### Association Between Socio-Demographic Factors and Blood Pressure Control

Our study revealed that there was a significant association between age and level of education with blood pressure control. Uncontrolled blood pressure was significantly (p <0.001) higher at 73.5% among individuals aged 56-65 years old as compared with individuals who were aged 30-45 years old at 34.1%. Moreover, uncontrolled blood pressure was significantly (p = 0.039) higher among individuals who attained no formal education (75%) as compared to those who attained tertiary level of education (40%).

However, there was no clear significant (p > 0.05) association between gender, marital status, religion, employment status, residence with level of blood pressure.

**Table 4:**
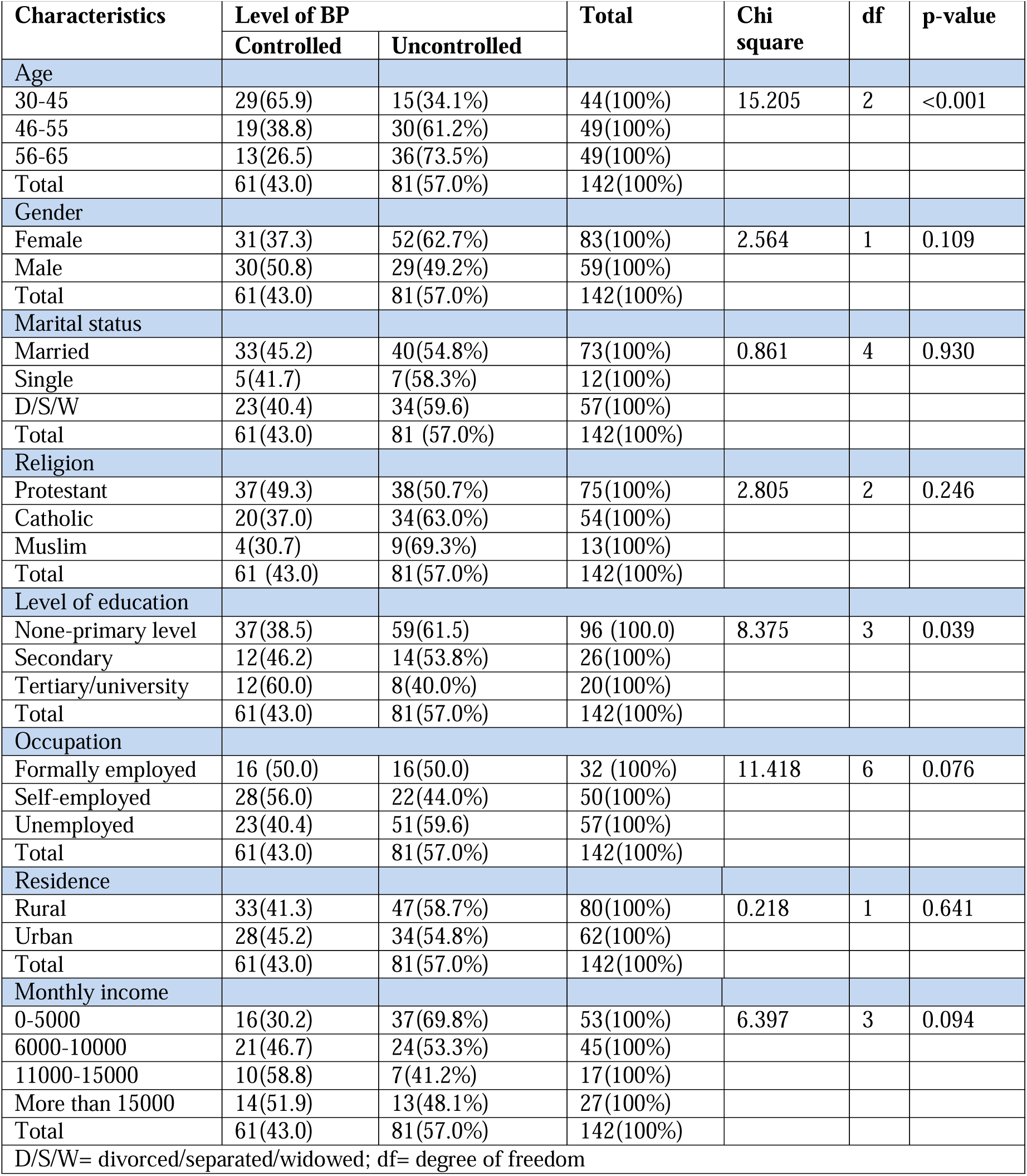
Association Between Socio-Demographic Factors and Blood Pressure Control (n, %)

### 4.6 Association Between Accessibility to Healthcare and Blood Pressure Control

Our study revealed a significant association between the frequency of visits to the Marung’a County Referral Hospital, affordability of the services provided, and access to most medical professionals in time, with the participant’s blood pressure control. Uncontrolled BP was also significantly (p=0.003) higher among respondents who rarely visited the Referral Hospital at 70.1% as compared to those who visited the hospital frequently at 45.3%.

Uncontrolled blood pressure was significantly (p<0.001) higher among the respondents who reported unaffordability of the healthcare services at 69.1% compared to those who reported affordability at 31.1%. Moreover, uncontrolled BP was significantly (p=0.013) higher among the respondents who reported not accessing the medical professionals in time at 65.9% in full comparison to those who reported accessing the medical professionals in good time at 45.0%.

However, there was no significant (p>0.05) association between distance from the hospital, type of road to the hospital, means of transport to the hospital, affordability of the means of transport, and how the respondents paid their medical bills with blood pressure control. Also, the Non-association between paying medical bills with the control of blood pressure was striking and equally intriguing to us, calling for further study.

**Table 5:** Association Between Accessibility to Healthcare and Blood Pressure Control (n,%)

| Characteristics | Level of BP |  | Total | Chi square | df | p-value |
| --- | --- | --- | --- | --- | --- | --- |
|  | Controlled | Uncontrolled |  |  |  |  |
| Distance from the hospital |  |  |  |  |  |  |
| Less than 1km | 25(49.0%) | 26(50.0%) | 51(100%) | 1.94 | 2 | 0.551 |
| 1-3 km | 21(39.6%) | 32(60.4%) | 53(100%) |  |  |  |
| More than 3km | 15(39.5%) | 23(60.5%) | 38(100%) |  |  |  |
| Total | 61(43.0%) | 81(57.0%) | 142(100%) |  |  |  |
| Type of road |  |  |  |  |  |  |
| Murram | 21(39.6%) | 32(60.4%) | 53(100%) | 0.384 | 1 | 0.536 |
| Tarmac | 40(44.9%) | 49(55.1%) | 89(100%) |  |  |  |
| Total | 61(43.0%) | 81(57.0%) | 142(100%) |  |  |  |
| Means of transport |  |  |  |  |  |  |
| Foot | 30(46.2%) | 35(53.8%) | 65(100%) | 0.942 | 2 | 0.624 |
| Private | 11(45.8%) | 13(54.2%) | 24(100%) |  |  |  |
| Public | 20(37.7%) | 33(62.3%) | 53(100%) |  |  |  |
| Total | 61(43.0%) | 81 (57.0%) | 142(100%) |  |  |  |
| Is the means of transport affordable |  |  |  |  |  |  |
| No | 15(44.1%) | 19(55.9%) | 34(100%) | 0.025 | 1 | 0.876 |
| Yes | 46(42.6%) | 62(57.4%) | 108(100%) |  |  |  |
| Total | 61(43.0%) | 81(57.0%) | 142(100%) |  |  |  |
| Frequency of visit to hospital |  |  |  |  |  |  |
| Once | 20(29.9%) | 47(70.1%) | 67(100%) | 8.893 | 1 | 0.003 |
| More than once | 41(54.7%) | 31(45.3%) | 75(100%) |  |  |  |
| Total | 61(43.0%) | 81(57.0%) | 142(100%) |  |  |  |
| Affordability of healthcare services |  |  |  |  |  |  |
| No | 30(30.9%) | 67(69.1%) | 97(100%) | 18.077 | 1 | < 0.001 |
| Yes | 31(68.9%) | 14(31.1%) | 45(100%) |  |  |  |
| Total | 61(43.0%) | 81(57.0%) | 142(100%) |  |  |  |
| Access |  |  |  |  |  |  |
| No | 28(34.1%) | 54(65.9%) | 82(100%) | 6.149 | 1 | 0.013 |
| Yes | 33(55.0%) | 27(45.0%) | 60(100%) |  |  |  |
| Total | 61(43.0%) | 81(57.0%) | 142(100%) |  |  |  |
| Bills |  |  |  |  |  |  |
| Cash | 31(42.5%) | 42(57.5%) | 73(100%) | 0.015 | 1 | 0.903 |
| Insurance | 30(43.5%) | 39(56.5%) | 69(100%) |  |  |  |
| Total | 61(43.0%) | 81(57.0%) | 142(100%) |  |  |  |

df=degree of freedom

### 4.7 Association Between Availability of Social; Support Network and BP Control

We found there was a significant association between having friends and family visit when sick, having emotional support, having financial support, and discussing health related problems with blood pressure control. Uncontrolled BP was significantly (p <0.001) higher among individuals who reported never visited by friends or family when they are sick (88.2%) compared to those who reported being visited by friends and family when they are sick (47.2%). Respondents who did not have emotional support was significantly (p=0.048) more likely to have uncontrolled BP at 71.4% compared to those who had emotional support at 52.0%.

Unregulated blood pressure was significantly (p=0.003) higher among respondents who reported not having financial support (74.0%) in comparison to those who had financial support (47.0%). Moreover, uncontrolled blood pressure was significant (p=0.010) higher among respondents who reported not having friends or family to discuss health-related problems at 71.2% as compared to those who had friends or family to discuss health-related problems at 48.9%.

**Table 6:** Association Between Availability of Social Support Network and Blood Pressure Control (n, %).

| Characteristics | Level of BP |  | Total | Chi square | df | p-value |
| --- | --- | --- | --- | --- | --- | --- |
|  | Controlled | uncontrolled |  |  |  |  |
| When I am sick, my family and/or friends visit me |  |  |  |  |  |  |
| No | 4 (11.8%) | 30 (88.2%) | 34 (100%) | 17.751 | 1 | <0.001 |
| Yes | 57 (52.8%) | 51 (47.2%) | 108(100%) |  |  |  |
| Total | 61 (43.0%) | 81 (57.0%) | 142(100%) |  |  |  |
| I usually get emotional support from my family and/or friends |  |  |  |  |  |  |
| No | 10(28.6%) | 25 (71.4%) | 35(100%) | 3.923 | 1 | 0.048 |
| Yes | 51(47.7%) | 56(52.3%) | 107(100%) |  |  |  |
| Total | 61(43.0%) | 81(57.0%) | 142(100%) |  |  |  |
| I usually get financial support from my family and/or friends |  |  |  |  |  |  |
| No | 13(26.0%) | 37(74.0%) | 50(100%) | 9.057 | 1 | 0.003 |
| Yes | 48(52.2%) | 44(47.8%) | 92(100%) |  |  |  |
| Total | 61(43.0%) | 81(57.0%) | 142(100%) |  |  |  |
| When I face a problem, my family and/or friends help me solve it |  |  |  |  |  |  |
| No | 15(28.8%) | 37(71.2%) | 52(100%) | 6.668 | 1 | 0.010 |
| Yes | 46(51.1%) | 44(48.9%) | 90(100%) |  |  |  |
| Total | 61(43.0%) | 81(57.0%) | 142(100%) |  |  |  |

df= degree of freedom

## 5. DISCUSSION

Our study found that most (57%) of the respondents at the Murang’a County Referral Hospital in Kenya had uncontrolled blood pressure levels that were directly associated with vatious distinct Social Determinants of Health (SDOH), such as:

Advanced age,
Illiteracy,
Less frequency of hospital visits,
Lack of access to healthcare services
Lack of affordability of healthcare services,
Lack of a psychosocial support system.

The levels of uncontrolled blood pressure in our study are too far off to meet the voluntary global target of 21% (Kario et al., 2023), as recommended by WHO (2023). In line with this finding, another study performed in Indonesia by Mitra et al. (2019) reported an uncontrolled BP level of 52.6% among patients with hypertension aged 60 to 77 years.

### Age, Illiteracy and Frequency of Hospital Visit in Relation to BP Control

In our study, uncontrolled blood pressure was significantly higher among the patients with hypertension, aged 56 years and above. This finding agrees with other studies conducted in Kenya (Mohammed et al., 2018) and South Africa (Monakali et al., 2018) that showed advanced age as significant risk for uncontrolled BP. This finding is also like previous cohort studies that examined BP control in adults with HTN (Myers et al., 2024; Osude et al., 2021). A study carried out in the United States confirmed that advanced age and low education level as both significantly impacting blood pressure control (Lin et al., 2022). According to Elnaem et al. (2021), older age groups exhibited higher rates of uncontrolled blood pressure while those with higher education levels showed better control. The direct association between age and levels of BP might be related to the fact that adult aging processes attributed to changes such as weight gain, compromised important compensatory mechanisms, notably baroreceptor reflex, kidney’s buffering capacity, stiffness of blood vessels are associated with high blood pressure (Rinnstrom et al., 2017). These findings underscore the need for policymakers and healthcare providers to design and implement age-specific interventions by paying more attention to the elderly with hypertension.

Our study found that uncontrolled BP was substantially higher among individuals who had no formal education compared to those who had completed tertiary level of education. This finding correlates with studies conducted in Kenya (Gatimu & John, 2020) and China (Sun et al., 2022) that revealed uncontrolled BP was significantly higher among individuals with a primary level of education compared with those who attained higher levels of education. This might be related to the fact that people with higher education levels are more likely to be health conscious and are more likely to want to control their blood pressure levels. Choi & Kim (2023) and Hosseini et al. (2021) emphasized the importance of age and education level in shaping BP management plans and strategies. Also, these findings highlight the necessity for tailored interventions to address sociodemographic differences in the treatment of adult hypertension.

### Access to Healthcare Services in Relation to BP Control

Our study revealed that uncontrolled levels of blood pressure were considerably higher among respondents who could not afford and likewise access healthcare services. This finding concurs with a similar study conducted in Ghana (Adomako et al., 2021) that proved affordability of healthcare services and accessibility to antihypertensive drugs contribute to effective BP control. Another study also reported a significant correlation between patient ability to afford medications and adequate BP control (Harrison, 2021). The justification could be when there is prohibitive cost of common antihypertensive drugs, there is high chances of patients with HTN not purchasing these drugs which would lead to non -adherence to medication and poor BP control. Studies performed in Tanzania revealed that access to healthcare as a central factor in shaping management (Edward et al., 2021; Powell-Jackson et al., 2023) of blood pressure. Indeed, we see that these challenges in accessing healthcare services, including long road travel distances and limited availability of healthcare providers, hinder hypertension diagnosis and treatment (Dhungana et al., 2021).

We found that income inequality has profound effects on BP regulation, emphasizing the need to address Social Determinants of Health to reduce healthcare disparities (Gatimu & John, 2020). These findings demonstrate how important it is to address all aspects of the healthcare system to improve hypertension management and outcomes in setting with limited healthcare resources, especially in developing countries.

This study also established a significant association between uncontrolled blood pressure and frequency of visits to the hospital by the respondents. Uncontrolled adult BP was higher among respondents who rarely visited the hospital as compared to those who frequently visited the hospital for checkups. This finding is in line with a study conducted by Shima et al. (2016) that revealed patients who frequently visited the health facility are more likely to be health conscious and likely to follow the recommendations of their physicians to enhance lifestyle modification.

Frequent and brief intervals between hospital visits for hypertensive patients were linked to better control of their BP levels (King et al., 2017). When doubtful patients visit a healthcare facility infrequently, they fail to review their regimen arrangements and the patients are also unable to receive advice on non-pharmacological therapy, all which may contribute to uncontrolled BP

(Fentaw et al., 2022). This positive association between regularity in treatment follow-up and controlled BP could be because the regular treatment follow-up visits provide an opportunity for health care providers to educate patients about their disease and drug therapy, plus optimize and monitor treatment outcomes (Mahmood et al., 2020).

### Social Support Network in Relation to BP Control

According to our study, respondents who lack social support network were more likely to have uncontrolled blood pressure levels as compared to those who reported having a reliable social support system. Consistent with our findings, a study conducted in Nigeria showed that a high degree of family support was associated with better hypertension control (Ojo O. et al., 2016).

Our findings also concur with a study carried out by Harding et al. (2022) that showed emotional support was associated with controlled BP levels. Having friends and family visit when sick, helps in developing emotional and financial support, plus engagement in discussions related to health problems and decision making, all have a beneficial effect on blood pressure stability.

Psychosocial factors, components of Social Determinants of Health that induce emotional stress, can trigger a physiological reaction that is meditated by activation of the sympathetic nervous system. Repeated activation of this system can result in failing to return to resting BP levels (Spruill, 2010). Availability of a strong psychosocial support system significantly aids and helps hypertensive patients adhere to their prescribed regimen, a crucial factor to control their blood pressure (Gulcan et al., 2019). Additionally, social support can help moderate BP by reducing stress, improving self-esteem, and promoting good health behaviors.

### Strength and Limitation

Our hospital-based (Murang’a County Referral Hospital) cross-section research designs gives a clear view of the magnitude of the burden of blood pressure control. However, the sample utilized was small but representative, we believe, thus cautiously limits generalization of our data and findings to different contexts. The degree of influence of the Social Determinants of Health we examined with blood pressure control should be interpreted with caution and limited to a specific context.

## 6. CONCLUSION

Despite the availability of antihypertension medications, blood pressure control rates of the adult participants in our study were suboptimal. Advanced age, illiteracy, lack of access and scope of affordability of healthcare services, less frequency of hospital visits, and lack of a psychosocial support system were the social determinants of health associated with uncontrolled blood pressure levels.

Addressing these disparities through policy interventions and community-based strategies is crucial for improving overall health outcomes in settings with limited resources. These findings emphasize the need for tailored interventions that consider social determinants of health in HTN management. We suggest that integrating social determinants into clinical care such as community-based interventions can improve patient outcomes.

### STATEMENTS and DECLARATIONS CONSENT for PUBLICATION

All authors have acknowledged, agreed, and joined *in toto* for the publication of this research project and submission of the corresponding manuscript to the International Journal of Social Determinants of Health and Health Services (Sage Journal).

## ACKNOWLEDGEMENT

The authors gratefully acknowledge the untiring support of the research participants, healthcare workers, and hospital staff and officials who contributed to this study. Their participation and support were invaluable to our research, highly spirited, and equally motivational for all authors.

Also, we appreciate the encouragement and added insight received from lively discussions with our colleagues, students, and interested researchers.

## ETHICAL APPROVAL & INFORMED CONSENT STATEMENT

Ethical approval for our research study was sought and received from Murang’a County Referral Hospital and School of Nursing at The Catholic University of Eastern Africa. Our study protocol was approved by the Kenyatta National Hospital and the University of Nairobi.

An inform consent form was issued to each participant to confirm their understanding and assure them their contribution to the study will be confidential and all data provided will only be used for the purpose of the study. Respondents were also informed that no form of financial incentive will be given to them for being part of the study -- and that their choice of not to engage with the study cannot stop them from getting any benefit that may come because of the project..

## DATA AVAILABILITY

All data generated by this study are presented in the article, and the data are openly available to any interested person from the Lead Author (OTO) via a written or electronic media request.

## ABBREVIATIONS

MRRH: Murang’a Country Referral Hospital
SDOH: Social Determinants of health
HTN: Hypertension
BP: Blood Pressure
CVD: Cardiovascular Disease
SSA: Sub-Saharan Africa
LMIC: Low-and Medium-Income Countries
NCD: Non-Communicable Disease
KSh: Kenyan Schilling (currency)

## TRANSPARENCY

The authors volunteer that *ChatGPT* or any equivalent AI program was not a source of data or information and was not used to draft or embellish this manuscript.

## AUTHORS CONTRIBUTIONS

Okubastion Tekeste Okube Co-Conceptualization, Methodology Vincencia

Aketc Co-Conceptualization, Methodology, and Writing – review & editing.

Nerry Kitur Methodology and Writing – review & editing.

Josphat Njoroge Methodology and Writing – review & editing.

Norah Anne Mogute Oyagi Investigation, Formal Analysis, Validation,

and Writing – original draft.

Lee Presley Gary, Jr. Resources, Visualization, and Manuscript – reviewing and editing.

